# The Association between the Trajectory of the Life’s Essential 8 and Long-term Blood Pressure Variability: The Kailuan Study

**DOI:** 10.1101/2025.06.26.25330378

**Authors:** Shasha An, Zhao Han, Tairan Wang, Pei Liang, Weizhe Li, Xin Wang, Xueliang Ma, Yanxiu Wang, Kangran Li, Shouling Wu

**Affiliations:** Department of General Practice, Handan Central Hospital, Handan, Hebei, 056000, People’s Republic of China; Department of Dermatology, The Affiliated Hospital of Hebei Engineering University, Handan, Hebei, 056002, People’s Republic of China; Department of Severe Rehabilitation, Handan Mingren Hospital, Handan, Hebei, 056000, People’s Republic of China; Department of Emergency Internal Medicine I, Handan Central Hospital, Handan, Hebei, 056000, People’s Republic of China; Department of Cardiology, Kailuan General Hospital, Tangshan, Hebei, 063001, People’s Republic of China

**Keywords:** The trajectories of Life’s essential 8, Blood Pressure Variability, Prospective Cohort Study

## Abstract

This study aims to explore the relationship between the Trajectory of the Life’s Essential 8 and long-term blood pressure variability (BPV). A prospective cohort study was conducted using participants from Kailuan Group who participated in health examinations from 2006 to 2012, had no missing data in eight health indicators, and had complete baseline systolic and diastolic blood pressure data in 2006, as well as those with a history of myocardial infarction, cerebrovascular infarction, malignancy, or atrial fibrillation, were excluded. A total of 24 158 participants were included in the final cohort. We used four LE8 (Life’s Essential 8) scores from 2006 to 2012 to analyze their trajectories. To classify distinct trajectory groups within the study population, we employed latent class mixed models using the PROC TRAJ procedure in SAS. After comprehensive consideration, four distinct trajectories were identified. Subsequently, generalized linear regression models were applied to examine the association between LE8 score trajectories and blood pressure variability (BPV).

**Result:1:** The overall population’s SCV was **8**.42%, with the Very low-stable,Low-stable, the Median-stable and the High-stable trajectory showing SCVs of 8.83%, 8.53%, 8.34%, and 8.16%, respectively.There were statistically significant differences between the groups (P<0.05). The overall population’s DCV was 7.93%, with the Very low-stable, Low-stable, the Median-stable and the High-stable trajectory showing SCVs of 8.16%, 8.04%, 7.87%, and 7.70%, respectively.There were statistically significant differences between the groups (P<0.05). 2. The generalized Linear Regression of trajectories of LE8 and SCV, after adjusting for other risk factors, showed that compared to the Very low-stable trajectory, the SCV in the Low-stable, the Median-stable and the High-stable trajectory decreased by 0.227% (P <0.05), 0.392% (P < 0.05) and 0.347% (P < 0.05), respectively. It was also found that after adjusting for other risk factors, showed that compared to the Very low-stable trajectory, the DCV in the Low-stable, the Median-stable and the High-stable trajectory decreased by 0.193% (P >0.05), 0.332% (P < 0.05) and 0.431% (P < 0.05), respectively. Conclusion the trajectories of Life’s essential 8 is negatively correlated with long-term blood pressure variability.

---

A healthy lifestyle is the cornerstone for preventing cardiovascular diseases. In 2022, the American Heart Association (AHA) proposed 8 elements of cardiovascular health behaviors and factors[1,2], abbreviated as “Life’s Essential 8 (LE8)”. Relevant studies have confirmed that good cardiovascular health behaviors and factors have a protective effect on cardiovascular outcomes. The Framingham Heart Study found that compared with participants with LE8 < 68 (the sample median), those with LE8 ≥ 68 had a 53% lower risk of cardiovascular disease incidence and a 45% lower risk of death[3].

A meta-analysis study found that the group with high blood pressure variability (BPV) had a 67% increased risk of adverse cardiovascular events compared with the group with low BPV[4]. Long-term blood pressure variability (BPV) is influenced by multiple internal and external factors: physical inactivity, high body mass index (BMI), smoking, high-salt diet, etc., all of which can increase long-term BPV. We have previously found that the 7 indicators of cardiovascular health behaviors and factors promulgated by the AHA in 2010 were negatively correlated with long-term BPV. However, considering that health-related behaviors and factors are influenced by life changes, health status, or aging, we hypothesized that the trajectory score of LE8 is negatively correlated with long-term BPV by tracking how individual lifestyle and health indicators develop over time. To verify this hypothesis, we analyzed the association between the LE8 trajectory score and long-term blood pressure variability in the population of the Kailuan Study (ChiCTR-TNC-11001489) based on its dataset.

## 1. Materials and Methods

### 1.1 Study Population

The Kailuan Study is a prospective cohort study based in the Kailuan community of Tangshan. Tangshan is a large, modern city located along the Bohai Sea, while the Kailuan community is a comprehensive, well-managed area owned and administered by the Kailuan Group. Health services in this community are provided by 11 hospitals. Between 2006 and 2007, the first health examination was conducted for both active and retired employees of the Kailuan Group. Subsequently, every two years, health examinations are carried out by the same medical personnel at the same locations, following the same chronological order as the initial examination. The surveys include general information, individual health details, medication use, family health history, and lifestyle habits (including tobacco exposure, physical activity, salt intake, tea drinking habits, high-fat diet, sleep duration, and alcohol consumption), as detailed in previous studies[12]. Physical examinations include measurements of height, weight, and blood pressure, while biochemical indicators assessed include fasting plasma glucose (FBG), triglycerides (TG), total cholesterol (TC), high-density lipoprotein cholesterol (HDL-C), and low-density lipoprotein cholesterol (LDL-C), among others. The content of the survey, physical examination, and biochemical assessments remain consistent with the initial health examination. To date, a total of 8 follow-up surveys have been conducted. For further details regarding the Kailuan Study, please refer to previous literature[5-6].

### 1.2 Inclusion and Exclusion Criteria

Inclusion Criteria: (1)Participants completed physical examinations in the Kailuan Group during the following periods: 2006–2007, 2008–2009, 2010–2011, and 2012–2013, with complete data on cardiovascular health behaviors/factors and baseline blood pressure values; (2) Individuals who agreed to participate in this study and signed the informed consent form. Exclusion Criteria: (1) Individuals with a history of stroke (excluding lacunar infarction), myocardial infarction, malignant tumors, atrial fibrillation, or atrial flutter; (2) Participants with missing blood pressure data or extreme values in at least one of the 3 annual health check-ups in 2012 and subsequent years.

### 1.3 Data Collection

(1)Epidemiological Survey Form: The survey form will be completed by the participants. It will first be given to individuals for self-completion, and then, on the day of the health examination, the form will be verified for accuracy through face-to-face confirmation by medical personnel who have received standardized training. The form includes the following sections: General Information;Personal Health Information: Lifestyle habits, medication usage;Family Health History;Dietary and Lifestyle Habits: Smoking status, physical activity, salt intake, tea consumption, high-fat diet, sleep duration, alcohol consumption.Smoking Information: Includes current smoking status, age at which smoking started, and age at which smoking ceased. Physical Activity: Assessed by asking participants how much time they spent each week on vigorous, moderate, or light physical activities in the past 12 months. Sleep Duration: Measured by asking how many hours on average they sleep each night. Dietary Intake: Evaluated by asking about the frequency and portion size of typical foods consumed over the past year. Tea Drinking Habit: Includes categories such as never, once a month, or weekly tea consumption.High-Fat Diet: Categorized as rarely, occasionally, or frequently. Details are as previously described[7].

(2)Physical Examination: The physical examination includes the following measurements: height, weight, and blood pressure. All measurements will be carried out by trained healthcare personnel according to standardized protocols. Height and weight will be measured using a calibrated RGZ-120 model scale. The subjects should wear only light clothing, without shoes or hats. Height will be measured to the nearest 0.1 cm, and weight will be measured to the nearest 0.1 kg. Body Mass Index (BMI) will be calculated using the formula:BMI=Weight (kg)/Height (m)2. Blood Pressure Measurement: Blood pressure will be measured between 7:00 and 9:00 AM on the day of the health examination. Participants should refrain from smoking, drinking tea or coffee, or engaging in strenuous physical activity for 30 minutes prior to the measurement. The individual will sit quietly for 15 minutes with their back supported. The blood pressure will be measured while the subject is seated, with the arm at heart level, feet flat on the floor. A calibrated mercury sphygmomanometer will be used to measure the right brachial artery pressure. The cuff should be appropriately sized to cover at least 80% of the upper arm, with the lower edge of the cuff positioned 2-3 cm above the elbow and centered on the brachial artery. Systolic Blood Pressure (SBP): Measured at the first Korotkoff sound (Phase I). Diastolic Blood Pressure (DBP): Measured at the fifth Korotkoff sound (Phase V). Blood pressure will be recorded to the nearest 2 mmHg. Three measurements will be taken with a 1-2 minute interval between each, and the average of the three readings will be used. All measurements will be strictly conducted by trained personnel in accordance with the established standards. For further details, refer to previous literature[5-6].

(3)Laboratory Data Collection: Biochemical indicators to be tested include fasting plasma glucose (FBG), triglycerides (TG), total cholesterol (TC), high-density lipoprotein cholesterol (HDL-C), and low-density lipoprotein cholesterol (LDL-C). Non-high-density lipoprotein cholesterol (non-HDL-C) is calculated as: non-HDL-C=TC−HDL−C. After an 8-hour fast, blood will be drawn from the cubital vein (5 ml) on the morning of the examination. The sample will be collected into a vacuum tube and processed immediately. The blood will be centrifuged at 3000×g for 10 minutes at room temperature (24°C), and the serum will be separated. Measurement must be performed within 4 hours after sample collection.Glucose testing kit provided by Zhongsheng Beikou Biotechnology Co., Ltd.Reagents for TC, HDL-C, LDL-C, and TG provided by Shanghai Mingdian Bioengineering Co., Ltd. All tests will be conducted according to the reagent manufacturer’s instructions. As mentioned previously, measurements will be performed by trained personnel using standardized methods for blood pressure and blood sample collection^[8]^.

### 1.4 Relevant Definitions

#### 1.41 Measurement and quantification of LE8 score

Since the Kailuan study began in 2006, there has been a lack of detailed dietary data. Additionally, considering the impact of salt intake, tea consumption, and high-fat food intake on cardiovascular disease risks in the Chinese population, as previously mentioned, salt intake, tea intake, and high-fat food intake, based on questionnaire surveys, were used as alternative indicators of dietary quality [9-12].

According to the updated assessment method, the definition and scoring of the LE8 components include four health behaviors (diet, physical activity, smoking status, and sleep) and four health factors (BMI, non-HDL-C, blood glucose, and blood pressure). Each component of the LE8 is scored on a scale from 0 to 100. The total LE8 score is calculated by summing the scores of the eight indicators and dividing the total by 8. Therefore, the LE8 score ranges from 0 to 100. Based on the score, the LE8 levels are categorized into three groups: the Ideal LE8 group (80–100 points), the Intermediate LE8 group (50–79 points), and the Low LE8 group (0–49 points).

#### 1.42 Blood Pressure Variability (BPV)

BPV is represented by the coefficient of variation (CV) of blood pressure, which is calculated as the standard deviation of blood pressure divided by the mean blood pressure, multiplied by 100% (CV = SD/mean blood pressure×100%). The coefficient of variation for systolic blood pressure is referred to as SCV, and for diastolic blood pressure, it is referred to as DCV. The annual average and standard deviation (SD) of systolic blood pressure are calculated based on at least three years of systolic blood pressure values.

### 2. Statistical Methods

The data were entered through terminals at various health checkup hospitals and subsequently uploaded to the server in the computer room of the Kailuan General Hospital, forming an Oracle 10.2g database. Statistical analysis was conducted using SAS 9.4 software. For normally distributed continuous variables, the mean ± standard deviation (±s) was used. One-way analysis of variance (ANOVA) was applied for comparisons among more than two groups, with pairwise comparisons performed using the SNK method (for equal variances) or the Dunnett T3 method (for unequal variances). Non-normally distributed continuous variables were analyzed using the rank sum test and were expressed as interquartile ranges. Categorical data were expressed as n (%), with group comparisons made using the chi-squared (χ^2^) test. A latent mixture model within the PROC TRAJ procedure was employed to classify the trajectory patterns of LE8 from 2006 to 2012, following established methodology[13,14]. Various parameters and fit statistics were used to determine the optimal number of trajectories, including the proportion in each trajectory, Bayesian Information Criteria, average posterior probability in each trajectory, and visual inspection of the trajectories.The model with four trajectories provided the best fit. Models with different functional forms (e.g., 3-cubic, 2-quadratic, and 1-linear) were compared and evaluated based on their significance levels, utilizing the highest polynomial term. Ultimately, we identified four pattern with quadratic order terms, which met the statistical conditions necessary for constructing trajectory patterns. The Very low-stable trajectory was set as the reference for subsequent analysis. A generalized linear regression model was employed to analyze factors influencing long-term blood pressure variability (BPV), with a significance level of P < 0.05 (two-sided). The models used SCV and DCV as dependent variables, and LE8 grouping as the independent variable. Model 1 adjusted for age and sex; Model 2, based on Model 1, additionally adjusted for baseline systolic blood pressure; and Model 3 further adjusted for body mass index (BMI), triglycerides (TG), high-sensitivity C-reactive protein (hsCRP), alcohol consumption, and antihypertensive medication use. Considering the potential for multicollinearity among the independent variables, the variance inflation factor (VIF) was calculated. A VIF greater than 10 indicates multicollinearity. Variables exhibiting significant collinearity were excluded, and the generalized linear regression analysis was re-conducted.

## 3. Results

A total of 32,199 employees and retirees of Kailuan who participated in health examinations from 2006 to 2012 and had no missing data on eight health indicators, baseline systolic blood pressure, and diastolic blood pressure were included. 8,041 individuals met the exclusion criteria. Of these, 189 had a history of myocardial infarction, 147 had a history of stroke, 39 had a history of malignant tumors, and 119 had a history of atrial fibrillation or atrial flutter. Additionally, 1182 individuals had missing baseline blood pressure data. A total of 7,502 cases had fewer than 3 blood pressure measurements in health examinations after 2012, and 45 cases had extreme blood pressure values. Ultimately, 24,158 valid cases were included in the statistical analysis.

### 3.1 Characteristics of 24 158 participants according to the trajectories of Life’s essential 8 from 2006 to 2012

Among the 24,158 participants, the age ranged from 18 to 88 years (mean ± standard deviation: 46.14± 10.90). The distribution of the trajectories of LE8 was as follows: the Very low-stable group comprised 13.69% of the total population, the Low-stable group accounted for 84.81%, the Median-stable group accounted for 84.81%, and High-stable group represented 1.47%.

As the the trajectories of LE8 increased, there was a gradual decrease in the proportion of males, as well as in age, systolic blood pressure (SBP), diastolic blood pressure(DBP), body mass index (BMI), high-sensitivity C-reactive protein (hsCRP), triglyceride (TG) levels, and the proportion of individuals taking antihypertensive medications. However, alcohol consumption showed a gradual upward trend. These differences between groups were statistically significant (P < 0.05). (Table 1)

**Table 1:** Characteristics of 24 158 participants according to the trajectories of Life’s essential 8 from 2006 to 2012.

| Variables | LE8 scores trajectory patterns |  |  |  | P Value |
| --- | --- | --- | --- | --- | --- |
|  | Very low-stable group | Low-stable group | Median-stable group | High-stable group |  |
| Population, n (%) | 1847 | 8569 | 10152 | 3590 |  |
| AGE(years) | 45.69± 9.18 | 47.03± 10.39 | 47.76± 11.18 | 42.46±11.36 | <0.001 |
| MAN n (%) | 1778(96.3) | 7611(88.8) | 7684(75.7) | 1454(40.5) | <0.001 |
| SBP(mmHg) | 137.83±19.50 | 132.90±19.11 | 125.79±17.70 | 113.50±13.41 | <0.001 |
| DBP(mmHg) | 89.83±11.90 | 86.07±11.17 | 81.74±10.35 | 74.46±8.52 | <0.001 |
| BMI(kg/m<sup>2</sup>) | 27.76±3.32 | 26.24±3.26 | 24.61±3.14 | 22.39±2.69 | <0.001 |
| TG(mmol/L) | 1.99(1.32,3.13) | 1.52(1.08,2.33) | 1.21(0.89,1.75) | 0.93(0.67,1.24) | <0.001 |
| hsCRP(mg/L) | 0.99(0.40,2.30) | 0.79(0.29,2.15) | 0.60(0.21,1.70) | 0.40(0.16,1.24) | <0.001 |
| Alcohol Consumptionn (%) | 1114(60.3) | 6573(76.7) | 9013(88.8) | 3455(96.2) | <0.001 |
| Use of Antihypertensive Medication n (%) | 328(17.8) | 809(9.44) | 432(4.26) | 29(0.81) | <0.001 |

LE8, Life's Essential 8; SBP, systolic blood pressure; DBP, diastolic blood pressure; BMI, body mass index; TG: triglycerides; hsCRP: high-sensitivity C-reactive protein; 1 mmHg=0.133 kPa.

### 3.2 nDistribution of CV of 24 158 participants according to the trajectories of Life’s essential 8 from 2006 to 2012

In the total population, the SCV and DCV were 8.42% and 7.93%, respectively. As the LE8 score increased, SCV and DCV showed a gradual decreasing trend, with statistically significant differences between groups (P < 0.05). (Table 2).

**Table 2:** Distribution of CV of 24 158 participants according to the trajectories of Life’s essential 8 from 2006 to 2012.

| Variables | Very low-stable group | Low-stable group | Median-stable group | High-stable group | P Value |
| --- | --- | --- | --- | --- | --- |
| Participants, n (%) | 1847 | 8569 | 10152 | 3590 |  |
| SCV, (%) | 8.83(6.00,12.03) | 8.53(5.77,11.68) | 8.34(5.72,11.50) | 8.16(5.63,11.18) | <0.001 |
| DCV, (%) | 8.16(5.68,11.43) | 8.04(5.47,11.09) | 7.87(5.41,10.81) | 7.70(5.38,10.52) | <0.001 |

SCV, coefficient of the variation of systolic blood pressure; DCV, coefficient of the variation of diastolic blood pressure; LE8, Life’s Essential 8.

### 3.3 Generalized Linear Regression of trajectories of LE8 and SCV

We conducted a generalized linear regression analysis with SCV as the dependent variable and LE8 trajectory groups as the independent variable. In Model 1, after adjusting for age and gender, we found that compared to the Very low-stable trajectory, the SCV in the Low-stable, the Median-stable and the High-stable trajectory decreased by 0.326% (P < 0.05), 0.594% (P < 0.05) and 0.673% (P < 0.05), respectively. Model 2, built upon Model 1, further adjusted for baseline systolic blood pressure. It showed that compared to the Very low-stable trajectory, the SCV in the Low-stable, the Median-stable and the High-stable trajectory decreased by 0.254% (P < 0.05), 0.432% (P < 0.05) and 0.378% (P < 0.05), respectively. Model 3, built upon Model 2, further adjusted for age, BMI, TG, hsCRP, alcohol consumption, and antihypertensive medication use. It showed that for compared to the Very low-stable trajectory, the SCV in the Low-stable, the Median-stable and the High-stable trajectory decreased by 0.227% (P <0.05), 0.392% (P < 0.05) and 0.347% (P < 0.05), respectively (Table 3).

**Table 3:** Generalized Linear Regression of trajectories of LE8 and SCV.

|  | Very low-stable group | Low-stable group | Median-stable group | High-stable group |
| --- | --- | --- | --- | --- |
| Model 1 β(95%CI) | 1.000 (ref) | -0.326 (-0.548, -0.104) | -0.594(-0.815,-0.373) | -0.673(-0.934, -0.411) |
| Model 2 β(95% CI) | 1.000 (ref) | -0.254 (-0.477, -0.032) | -0.432(-0.657,-0.208) | -0.378 (-0.649, -0.106) |
| Model 3 β(95% CI) | 1.000 (ref) | -0.227(-0.454, -0.0002) | -0.392(-0.631,-0.154) | -0.347 (-0.643, -0.051) |

Note: p Values<0.05 for all statistical analyses in this table. Abbreviations: LE8, life's essential 8; SCV, coefficient of the variation of systolic blood pressure Model 1, adjusted for age and gender. Model 2, adjusted for baseline SBP on the basis of Model 1. Model 3, adjusted for BMI, TG, hsCRP, alcohol consumption and proportion of antihypertensive drug usage on the basis of Model 2.

### 3.4 Generalized Linear Regression of trajectories of LE8 and DCV

We conducted a generalized linear regression analysis with DCV as the dependent variable and LE8 trajectory groups as the independent variable. In Model 1, after adjusting for age and gender, we found that compared to the Very low-stable trajectory, the DCV in the Low-stable, the Median-stable and the High-stable trajectory decreased by 0.234% (P < 0.05), 0.436% (P < 0.05) and 0.687% (P < 0.05), respectively. Model 2, built upon Model 1, further adjusted for baseline systolic blood pressure. It showed that compared to the Very low-stable trajectory, the DCV in the Low-stable, the Median-stable and the High-stable trajectory decreased by 0.262% (P < 0.05), 0.454% (P < 0.05) and 0.609% (P < 0.05), respectively.

Model 3, built upon Model 2, further adjusted for age, BMI, TG, hsCRP, alcohol consumption, and antihypertensive medication use. It showed that for compared to the Very low-stable trajectory, the DCV in the Low-stable, the Median-stable and the High-stable trajectory decreased by 0.193% (P >0.05), 0.332% (P < 0.05) and 0.431% (P < 0.05), respectively (Table 4).

**Table 4:** Generalized Linear Regression of trajectories of LE8 and DCV.

|  | Very low-stable group | Low-stable group | Median-stable group | High-stable group |
| --- | --- | --- | --- | --- |
| Model 1 $\beta$(95%CI) | 1.000 (ref) | -0.234 (-0.449, -0.019) | -0.436(-0.648,-0.224) | -0.687(-0.926, -0.447) |
| Model 2 $\beta$(95% CI) | 1.000 (ref) | -0.262 (-0.477, -0.047) | -0.454(-0.671,-0.237) | -0.609(-0.871, -0.346) |
| Model 3 $\beta$(95% CI) | 1.000 (ref) | -0.193(-0.413, 0.025) | -0.332(-0.562,-0.103) | -0.431 (-0.717, -0.146) |

Note: p Values $< 0.05$ for all statistical analyses in this table.
Abbreviations: LE8, life's essential 8; DCV, coefficient of the variation of diastolic blood pressure
Model 1, adjusted for age and gender.
Model 2, adjusted for baseline DBP on the basis of Model 1.
Model 3, adjusted for BMI, TG, hsCRP, alcohol consumption and proportion of antihypertensive drug usage on the basis of Model 2.

### 3.5 Stratified analyses of Generalized Linear Regression of trajectories of LE8 and SCV

In further stratified analysis, There was an interaction between the group aged ≥60 years and the group aged <60 years. After adjusting for other confounders in young participants, compared with the Very low-stable trajectory group,SCV decreased by 0.330 in the High-stable group; However, this phenomenon was not observed in participants aged ≥60 years. There was no interaction between genders, but compared with the Very low-stable trajectory group, the SCV in the High-stable group decreased more significantly (0.496) among men. (Table 5).

**Table 5:** Stratified analyses of Generalized Linear Regression of trajectories of LE8 and SCV.

|  | Very low-stable group | Low-stable group | Median-stable group | High-stable group | P-interaction |
| --- | --- | --- | --- | --- | --- |
| Age | | | | | $< 0.001$ |
| <60 | 1.0(ref) | -0.241(-0.473,-0.010) | -0.378(-0.622,-0.134) | -0.330(-0.636,-0.024) |  |
| $\geq 60$ | 1.0(ref) | 0.389(-0.611,1.390) | 0.156(-0.867,1.179) | 0.735(-0.484,1.955) | |
| Gender | | | | | $> 0.05$ |
| Women | 1.0(ref) | 0.017(-1.103,1.138) | 0.026(-1.097,1.148) | 0.330(-0.841,1.502) |  |
| Men | 1.0(ref) | -0.250(-0.481,-0.019) | -0.442(-0.688,-0.198) | -0.496(-0.831,-0.161) |  |

Note: p Values $< 0.05$ for all statistical analyses in this table.
Abbreviations: LE8, life's essential 8; SCV, coefficient of the variation of diastolic blood pressure
Adjusted for age and gender, baseline SBP, BMI, TG, hsCRP, alcohol consumption and proportion of antihypertensive drug usage.

### 3.6 Stratified analyses of Generalized Linear Regression of trajectories of LE8 and DCV

In further stratified analysis, There was an interaction between the group aged ≥60 years and the group aged <60 years. After adjusting for other confounders in young participants, compared with the Very low-stable trajectory group,DCV decreased by 0.487 in the High-stable group; However, this phenomenon was not observed in participants aged ≥60 years. There was no interaction between genders, but compared with the Very low-stable trajectory group, the DCV in the High-stable group decreased more significantly (0.321) among men. (Table 6).

**Table 6:**
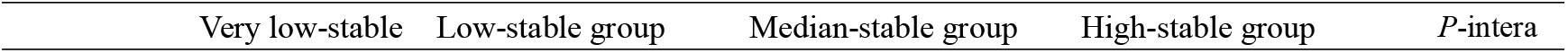
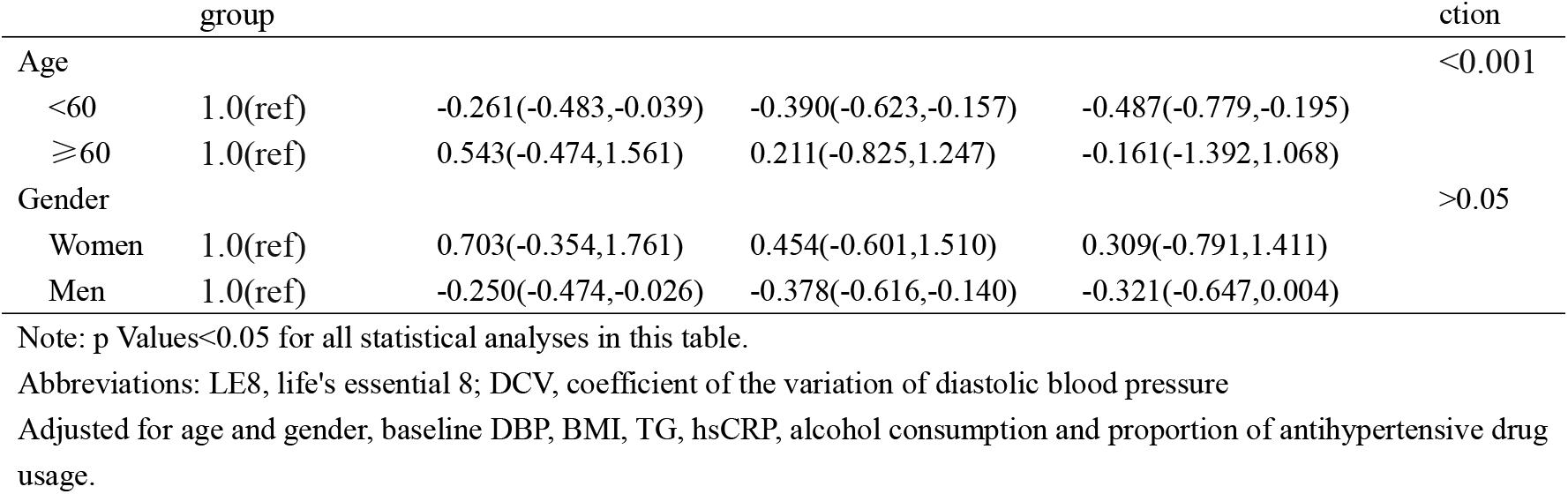
Stratified analyses of Generalized Linear Regression of trajectories of LE8 and DCV.

## Discussion

In this large prospective cohort study, we found that the trajectory of LE8 scores was negatively associated with long-term systolic blood pressure variability (SBPV), and this association was independent of baseline LE8 scores and other factors influencing long-term SBPV. Additionally, this association was also negatively correlated with long-term diastolic blood pressure variability. These results confirmed our hypothesis.

We observed that compared with the 6-year extremely low stable LE8 score trajectory, the lower stable LE8 score trajectory, moderate stable LE8 score trajectory, and increasing stable LE8 score trajectory were negatively associated with long-term SBPV, with β values of -0.227% (P < 0.05), -0.392% (P < 0.05), and -0.347% (P < 0.05), respectively; Similarly, the lower stable LE8 score trajectory, moderate stable LE8 score trajectory, and increasing stable LE8 score trajectory were negatively associated with long-term DBPV, with β values of -0.193% (P > 0.05), -0.332% (P < 0.05), and -0.431% (P < 0.05), respectively. Additionally, in the stratified analysis, we found that participants aged <60 years showed more significant reductions in systolic blood pressure variability (SCV) and diastolic blood pressure variability (DCV) under the increasing stable LE8 score trajectory, suggesting they may derive greater benefits.

Trajectory modeling captures the changes of variables or behaviors over time, It provides a dynamic and more accurate alternative to single measurements that only capture data at specific time points[15,16]. Since LE8-related factors can be influenced by life transitions, health status, or aging,Therefore, trajectory modeling is particularly beneficial for evaluating lifestyle metrics such as LE8.Therefore, the method of trajectory modeling is particularly advantageous for the evaluation of lifestyle metrics such as LE8[17].

There is still limited research on LE8 and BPV; Our previous research has found that LS7 is negatively correlated with systolic blood pressure variability[18]. However, a large number of literatures have studied the relationship between LE8 and arteriosclerosis as well as carotid artery plaques.Studies like those conducted by Liu Q et al. have found that the LE8 trajectory score is negatively correlated with carotid intimal thickening[19]; Other studies have found that the LE8 score is negatively correlated with carotid intimal thickening and ePWV[20-24].

Although there are few specific studies on LE8 and long-term BPV, but elements within the LE8 framework have been confirmed to be associated with BPV. Studies have found that a certain amount of exercise[25] and appropriate sleep[26] can reduce blood pressure variability; Other studies have found that smoking[27], high sodium chloride intake[28], elevated systolic blood pressure[29], poor blood glucose[29] and lipid profiles[29], and increased BMI[30] are positively correlated with systolic blood pressure variability.Since humans as a whole may simultaneously have multiple protective or risk factors, therefore, a comprehensive evaluation of the effects of these factors on long-term systolic blood pressure variability (SBPV) may be more scientific.

The main advantage of this study lies in its innovative examination of the relationship between LE8 score trajectories and BPV, and it is a prospective design, community-based large sample, and long follow-up period. However, our study has certain limitations: First, the 6-year duration of the LE8 trajectory in the current study may not fully reflect an individual’s lifetime cardiovascular health; Second, the current results rely heavily on questionnaires, which may introduce bias and reduce reliability; Third, our observation subjects were mainly male, resulting in an unbalanced gender ratio.

## data availability statement

This is an open access article under the terms of the Creative Commons Attribution-NonCommercial License, which permits use, distribution and reproduction in any medium, provided the original work is properly cited and is not used for commercial purposes.

## Author contributions

Concept and design: S. Wu, An, J. Gao, Y. Wu, Jin, X. Gao.

Acquisition, analysis, or interpretation of data: S. Wu, An, Li, Lichtenstein, Kris-Etherton, Jin, Huang, Hu, X. Gao.

Drafting of the manuscript: Shouling Wu, Shasha An.

Critical revision of the manuscript for important intellectual content: S. Wu, Shasha An, Tairan Wang.

Statistical analysis: S. Wu, Shasha An, Weizhe Li, Weizhe Li.

Administrative, technical, or material support: Pei Liang, Zhao Han.

Supervision: S. Wu, Tairan Wang.

## References

1. Grandner MA, Fernandez FX. The translational neuroscience of sleep: a contextual framework. Sci.2021;374(6567):568–73. doi:10.1126/science.abj8188.

2. Lloyd-Jones DM, Allen NB, Anderson CAM, Black, T, Brewer, LC, Foraker, RE, Grandner, MA, Lavretsky, H, Perak, AM, Sharma, G, et al. Status of cardiovascular health in US adults and children using the American Heart Association’s new “Life’s Essential 8” metrics: prevalence estimates from the National Health and Nutrition Examination Survey (NHANES), 2013-2018, Circulation, 2022;146(11):822–835.

3. Rempakos A, Prescott B, Mitchell GF, Vasan RS, Xanthakis V. Association of Life’s Essential 8 With Cardiovascular Disease and Mortality: The Framingham Heart Study. J Am Heart Assoc. 2023 Dec 5;12(23):e030764. doi: 10.1161/JAHA.123.030764. Epub 2023 Nov 28. PMID: 38014669; PMCID: PMC10727315.

4. Wang J, Shi X, Ma C, Zheng H, Xiao J, Bian H, Ma Z, Gong L. Visit-to-visit blood pressure variability is a risk factor for all-cause mortality and cardiovascular disease: a systematic review and meta-analysis. J Hypertens. 2017 Jan;35(1):10–17. doi: 10.1097/HJH.0000000000001159. PMID: 27906836.

5. Wu S, Huang Z, Yang X, Zhou Y, Wang A, Chen L, et al. Prevalence of ideal cardiovascular health and its relationship with the 4-year cardiovascular events in a northern Chinese industrial city. Circ Cardiovasc Qual Outcomes (2012) 5(4):487–93. doi: 10.1161/CIRCOUTCOMES.111.963694

6. Zheng X, Zhang R, Liu X, Zhao H, Liu H, Gao J, et al. Association between cumulative exposure to ideal cardiovascular health and arterial stiffness. Atherosclerosis (2017) 260:56–62. doi: 10.1016/j.atherosclerosis.2017.03.018.

7. Wu S, An S, Li W, Lichtenstein AH, Gao J, Kris-Etherton PM, et al. Association of trajectory of cardiovascular health score and incident cardiovascular disease. JAMA Netw Open (2019) 2(5):e194758. doi: 10.1001/jamanetworkopen.2019.4758

8. Jin C, Chen S, Vaidya A, Wu Y, Wu Z, Hu FB, et al. Longitudinal change in fasting blood glucose and myocardial infarction risk in a population without diabetes. Diabetes Care (2017) 40(11):1565–72. doi: 10.2337/dc17-0610.

9. Li Y, Huang Z, Jin C, Xing A, Liu Y, Huangfu C, et al. Longitudinal change of perceived salt intake and stroke risk in a Chinese population. Stroke (2018) 49(6):1332–9. doi: 10.1161/STROKEAHA.117.020277.

10. Zhang Q, Zhou Y, Gao X, Wang C, Zhang S, Wang A, et al. Ideal cardiovascular health metrics and the risks of ischemic and intracerebral hemorrhagic stroke. Stroke (2013) 44(9):2451–6. doi: 10.1161/STROKEAHA.113.678839.

11. Huang S, Li J, Wu Y, Ranjbar S, Xing A, Zhao H, et al. Tea consumption and longitudinal change in high-density lipoprotein cholesterol concentration in Chinese adults. J Am Heart Assoc (2018) 7(13):e008814. doi: 10.1161/JAHA.118.008814.

12. Wang X, Liu F, Li J, Yang X, Chen J, Cao J, et al. Tea consumption and the risk of atherosclerotic cardiovascular disease and all-cause mortality: the China-PAR project. Eur J Prev Cardiol (2020) 27(18):1956–63. doi: 10.1177/2047487319894685.

13. Jones BL, Nagin DS. A SAS Procedure based on mixture models for estimat ing Developmental trajectories. Sociol Methods Res. 2001;29:374–93.

14. Jones BL, Nagin DS. Advances in Group-based trajectory modeling and an SAS Procedure for estimating them. Sociol Methods Res. 2007;35:542–71.

15. Liu, T. et al. C-reactive protein trajectories and the risk of all cancer types: a prospective cohort study. Int. J. Cancer. 151 (2), 297–307 (2022).

16. Deng, L. et al. The association of metabolic syndrome scores trajectory patterns with risk of all cancer types. Cancer 130 (12), 2150–2159 (2024).

17. Mésidor, M., Rousseau, M. C., O’Loughlin, J. & Sylvestre, M. P. Does group-based trajectory modeling estimate spurious trajectories? BMC Med. Res. Methodol. 22 (1), 194 (2022).

18. An S, Bao M, Wang Y, Li Z, Zhang W, Chen S, Li J, Yang X, Wu S, Cai J. Relationship between cardiovascular health score and year-to-year blood pressure variability in China: a prospective cohort study. BMJ Open. 2015 Oct 26;5(10):e008730. doi: 10.1136/bmjopen-2015-008730. PMID: 26503389; PMCID: PMC4636657.

19. Liu Q, Cui H, Chen S, Zhang D, Huang W, Wu S. Association of baseline Life’s Essential 8 score and trajectories with carotid intima-media thickness. Front Endocrinol (Lausanne). 2023 Jun 2;14:1186880. doi: 10.3389/fendo.2023.1186880. PMID: 37334294; PMCID: PMC10272710.

20. Chavoshi V, Barzin M, Ebadinejad A, Dehghan P, Momeni Moghaddam A, Mahdavi M, Hadaegh F, Niroomand M, Valizadeh M, Azizi F, Mirmiran P, Hosseinpanah F. Association of ideal cardiovascular health with carotid intima-media thickness (cIMT) in a young adult population. Sci Rep. 2022 Jun 16;12(1):10056. doi: 10.1038/s41598-022-13994-5. PMID: 35710831; PMCID: PMC9203712.

21. Santos IS, Goulart AC, Pereira AC, Lotufo PA, Benseñor IM. Association between Cardiovascular Health Score and Carotid Intima-Media Thickness: Cross-Sectional Analysis of the Brazilian Longitudinal Study of Adult Health (ELSA-Brasil) Baseline Assessment. J Am Soc Echocardiogr. 2016 Dec;29(12):1207-1216.e4. doi: 10.1016/j.echo.2016.09.001.Epub 2016 Oct 20. PMID: 27773521.

22. Fan Y, Yang S, Ruan L, Zhang C, Cao M. Association between Life’s Essential 8 and estimated pulse wave velocity among adults in the US: a cross-sectional study of NHANES 2011-2018. Front Public Health. 2024 May 30;12:1388424. doi: 10.3389/fpubh.2024.1388424. PMID: 38873301; PMCID: PMC11169870.

23. Kaneko F, Lee H, Shim JS, Kim HC. Maintaining optimal cardiovascular health metrics and carotid intima-media thickness among Korean adolescents. Clin Hypertens. 2025 May 1;31:e16. doi: 10.5646/ch.2025.31.e16. PMID: 40336508; PMCID: PMC12055496.

24. Guo F, Chen X, Howland S, Maldonado LE, Powell S, Gauderman WJ, McConnell R, Yan M, Whitfield L, Li Y, Bastain TM, Breton CV, Hodis HN, Farzan SF. Association Between Cardiovascular Health and Subclinical Atherosclerosis Among Young Adults Using the American Heart Association’s “Life’s Essential 8” Metrics. J Am Heart Assoc. 2024 Aug 6;13(15):e033990. doi: 10.1161/JAHA.123.033990. Epub 2024 Jul 30. PMID: 39077816; PMCID: PMC11964082.

25. Xu X, Meng X, Oka SI. Long-Term Habitual Vigorous Physical Activity Is Associated With Lower Visit-to-Visit Systolic Blood Pressure Variability: Insights From the SPRINT Trial. Am J Hypertens. 2021 May 22;34(5):463–466. doi: 10.1093/ajh/hpaa198. PMID: 33245323.

26. Liu X, Logan J, Kwon Y, Lobo JM, Kang H, Sohn MW. Visit-to-visit blood pressure variability and sleep architecture. J Clin Hypertens (Greenwich). 2021 Feb;23(2):323–330. doi: 10.1111/jch.14162. Epub 2021 Jan 25. PMID: 33492762; PMCID: PMC8030048.

27. Ushigome E, Fukui M, Hamaguchi M, Tanaka T, Atsuta H, Mogami SI, Oda Y, Yamazaki M, Hasegawa G, Nakamura N. Factors affecting variability in home blood pressure in patients with type 2 diabetes: post hoc analysis of a cross-sectional multicenter study. J Hum Hypertens. 2014 Oct;28(10):594–9. doi: 10.1038/jhh.2014.2. Epub 2014 Feb 6. PMID: 24500720.

28. Simmonds SS, Lay J, Stocker SD. Dietary salt intake exaggerates sympathetic reflexes and increases blood pressure variability in normotensive rats. Hypertension. 2014 Sep;64(3):583–9. doi: 10.1161/HYPERTENSIONAHA.114.03250. Epub 2014 Jun 9. PMID: 24914195; PMCID: PMC4267536.

29. Shin JH, Shin J, Kim BK, Lim YH, Park HC, Choi SI, Kim SG, Kim JH. Within-visit blood pressure variability: relevant factors in the general population. J Hum Hypertens. 2013 May;27(5):328–34. doi: 10.1038/jhh.2012.39. Epub 2012 Sep 13. PMID: 22971753.

30. Chen H, Zhang R, Zheng Q, Yan X, Wu S, Chen Y. Impact of body mass index on long-term blood pressure variability: a cross-sectional study in a cohort of Chinese adults. BMC Public Health. 2018 Oct 22;18(1):1193. doi: 10.1186/s12889-018-6083-4. PMID: 30348124; PMCID: PMC6196453.

